# Toward Personalized Diabetic Retinopathy Screening: Deep Learning Fundus Image Analysis and Clinical Risk Factors

**DOI:** 10.64898/2026.07.28.26359113

**Authors:** Flavio Ragni, Paolo Bresolin, Madalina Cocu, Stefano Bovo, Giulia Malfatti, Diego Cagol, Sandro Inchiostro, Federica Romanelli, Monica Moroni, Giuseppe Jurman

## Abstract

Diabetic retinopathy (DR) screening commonly relies on fixed follow-up intervals, although progression risk differs across patients. We developed a preliminary image–clinical framework to support personalized follow-up recommendations from retinal fundus images and systemic risk factors. Public DR datasets were harmonized into a binary task distinguishing absence of DR from DR of any grade. An ImageNet-pretrained ResNet50 and a foundation model-based feature extraction pipeline were compared. The selected image model was integrated with literature-derived severe retinopathy progression curves and clinical risk modifiers to estimate personalized cumulative risk and assign follow-up intervals using a predefined acceptable risk threshold. The ResNet50-based model was selected for subsequent analyses. In the target cohort, the integrated model assigned 90.0% of patients to follow-up within 12 months. Clinical adjustment substantially modified image-only recommendations, generally shifting patients toward shorter intervals. These preliminary findings support the feasibility of combining image-derived estimates of baseline DR status with clinical modifiers to inform personalized screening intervals. Larger longitudinal studies are needed to validate calibration, clinical utility, and safety before real-world implementation.

## 1 Introduction

Diabetes is one of the most prevalent chronic diseases worldwide, affecting approximately 589 million people in 2024, and represents a major public health challenge. Diabetic retinopathy (DR) is among its leading causes of vision impairment and blindness, affecting roughly one in four people with diabetes over their lifetime [1]. Screening guidelines recommend regular fundus examinations for early detection and timely treatment, but the growing number of patients and limited ophthalmology resources negatively affect adherence to screening programmes. Large public retinal image datasets have enabled highly effective deep learning (DL) models for DR detection, with performance comparable to expert graders [2]. More recently, foundation models trained on millions of fundus images have become available [3]. However, when moving from cross-sectional classification to longitudinal prediction and multimodal integration, public datasets become much scarcer, limiting the development of robust data-driven models for progression over time. While statistical and epidemiological models have been used to characterize longitudinal DR progression [4], this work aims to integrate data-driven image analysis with established clinical risk factors to assess whether combining these complementary sources improves progression modeling over time.

## 2 Data and Methods

We propose a multimodal method combining retinal fundus images and clinical information to personalize screening intervals in diabetic patients. The method first estimates the probability of baseline DR presence from retinal images and then combines this information with clinical data and epidemiological knowledge to estimate personalized risk of progression to proliferative diabetic retinopathy (PDR) and corresponding screening intervals.

### 2.1 Datasets

We used the following public datasets for DR classification: Messidor2 [5, 6](N=1,748), IDRiD [7] (N=516), Takahashi et al. [8] (N=9,939), DDR [9] (N=13,673), Benitez et al. [10] version V0.3 (N=1,437), and APTOS 2019 [11] (N=3,662). Together, they include images acquired with more than 40 fundus cameras across hospitals in France, India, Japan, China, and Paraguay, providing a heterogeneous development and validation setting. We also included a private INSPIRED dataset of retinal fundus images and clinical data from diabetic patients in the Autonomous Province of Trento between 2024 and 2026 (N=30, 91 images). Clinical variables were selected from established DR risk factors and extracted from electronic healthcare records and patient questionnaires. The dataset was cleaned, standardized, and filtered to retain variables with high availability, while fundus images were labelled for DR by expert ophthalmologists.

### 2.2 DR Risk Prediction from Fundus Oculi Images

Only labelled public images were retained for binary DR classification, distinguishing absence of DR (class 0) from DR of any grade (class 1). Labelled images from Messidor2, IDRiD, Takahashi, and DDR were merged into a dataset of 25,872 images, with 54.16% negative samples. Data were split into training, validation, and internal test sets using an 80/15/5 split, preserving class distribution and applying patient-level separation when identifiers were available. Models were trained on the training set, optimized on the validation set and evaluated on the internal test set and three external cohorts: Benitez V0.3, APTOS 2019, and INSPIRED. Two model families were compared: pretrained Convolutional Neural Networks (CNNs) and vision foundation models. For the CNN approach, transfer learning was performed using a ResNet50 pretrained on ImageNet-1k, with additional fully connected layers and partial unfreezing of the last trainable blocks. Images underwent standard ResNet50 preprocessing, while CLAHE, Gaussian blur, random crops, horizontal flips, and color jitter were tested as tunable preprocessing or augmentation options. These were optimized together with the number of added fully connected layers, unfrozen blocks, learning rate, weight decay, early stopping patience, batch size, and dropout probability. The optimal configuration was selected by the lowest validation Binary Cross Entropy (BCE) loss. In parallel, RETFound [3] was used to extract 1024-dimensional embeddings from fundus images, which were classified through a three-layer fully connected network. The same train, validation, and test partitions were used for direct comparison with the CNN approach. A grid search optimized batch size, learning rate, hidden layer dimensions, and dropout probabilities, again selecting the best model by validation BCE loss. Final performance was assessed using accuracy, precision, recall, F1-score, and AUC-ROC on the internal and external test sets. After selecting the optimal model, probability calibration was applied to better align model outputs with the INSPIRED target population.

### 2.3 PDR Risk Curve Modeling and Visit Interval Recommendation

The probabilities estimated by the DR classification model were combined with literature-derived hazard ratios or risk ratios to generate a personalized PDR risk profile. Standard cumulative PDR progression curves were first constructed at fixed horizons from 6 to 60 months using published epidemiological data. For eyes without DR, 1-, 3-, and 5-year cumulative risks were taken from [12]. For eyes with baseline DR, an overall “any DR” curve was derived by weighting severity-specific PDR risks according to the observed prevalence of each grade:

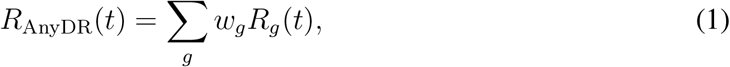

where *R*_*g*_(*t*) is the cumulative PDR risk for severity group *g*, and *w*_*g*_ is its prevalence weight. Weibull models were then fitted to the no-DR and any-DR profiles to obtain continuous progression curves.

For each eye, an expected image-based risk curve was computed as the probability-weighted combination of the two Weibull curves, avoiding a hard classification threshold:

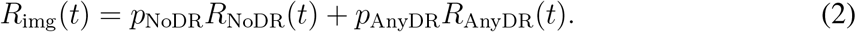

This risk was adjusted using a literature-informed clinical multiplier based on the selected predictors listed in Table 1.

**Table 1:** Selected clinical variables and corresponding effect sizes. Selected variables, corresponding hazard ratios or risk ratios, and reference studies used for the literature-informed clinical adjustment.

| Variable | HR/RR | Reference study |
| --- | --- | --- |
| Diabetes duration | < 10 years → RR = 1.38 (T1D); 1.00 (T2D)<br>10–20 years → RR = 2.43 (T1D); 2.06 (T2D)<br>20+ years → RR = 2.69 (T1D); 2.45 (T2D) | [13] |
| HbA1c (%) | RR = 1.29 per +1% HbA1c for progression | [14] |
| Systolic blood pressure | HR = 1.24 per +10 mmHg; baseline reference = 130 mmHg | [15] |
| Anaemia (T2D only) | $\log(1.29) \cdot I(\text{anaemia}) \cdot I(\text{T2D})$ | [16] |
| Fenofibrate | $\log(0.72) \cdot I(\text{fenofibrate yes})$ | [17] |
| Gender | Female RR = 2.74 (T1D); 1.78 (T2D)<br>Male RR = 3.30 (T1D, T2D) | [18, 19, 20] |

Log-relative risks or log-hazard ratios were summed and exponentiated to obtain the multiplicative clinical adjustment:

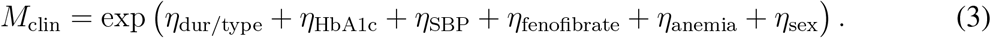

Under a proportional hazards assumption, the personalized cumulative risk curve was then computed as:

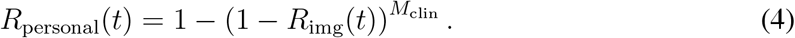

Given a predefined acceptable PDR risk threshold, i.e. 3%, before the next visit, the recommended follow-up interval was defined as the latest available time point preceding the first horizon at which *R*_personal_(*t*) exceeded that threshold. Because the clinical effect sizes were extracted from heterogeneous literature sources rather than jointly estimated in the target cohort, the integrated clinical layer should be interpreted as a literature-informed heuristic component within a blueprint framework rather than as a validated prognostic model.

## 3 Results

For binary classification of retinal fundus images, the best ResNet50 and foundation-model configurations were selected using validation BCE. The optimal ResNet50 model unfroze all pretrained blocks and added one fully connected layer; no additional preprocessing or augmentation improved performance beyond standard pretrained-image transformations. Final hyperparameters were a learning rate of 2e-4, weight decay of 6e-6, batch size of 32, and dropout of 0.5. The best foundation-model classifier used a hidden dimension of 256, dropout of 0.6, batch size of 16, and learning rate of 5e-5. After comparison on the internal test set and three external cohorts (Table 2), the ResNet50-based model was selected.

**Table 2:** Performance of fundus image classification models. Performances on test data of the best fundus image classification models across internal and external test datasets.

| Dataset | Architecture | Accuracy | Precision | Recall | F1-Score | AUC-ROC |
| --- | --- | --- | --- | --- | --- | --- |
| Internal Test | ResNet50 | **0.8613** | **0.8600** | **0.8255** | **0.8424** | **0.9330** |
|  | RETFound | 0.8245 | 0.8501 | 0.7504 | 0.7934 | 0.8976 |
| Benitez V0.3 | ResNet50 | **0.8894** | **0.8428** | **0.9601** | **0.8976** | **0.9689** |
|  | RETFound | 0.7300 | 0.6690 | 0.9215 | 0.7752 | 0.8874 |
| APTOS 2019 | ResNet50 | **0.7859** | **0.7033** | **0.9995** | **0.8256** | **0.9971** |
|  | RETFound | 0.7537 | 0.6793 | 0.9742 | 0.8004 | 0.9291 |
| INSPIRED | ResNet50 | **0.6044** | **0.2000** | **1.0000** | **0.3333** | **0.9444** |
|  | RETFound | 0.4835 | 0.1607 | 1.0000 | 0.2769 | 0.6856 |

The integrated image–clinical model mostly recommended short follow-up intervals: 8/30 patients were assigned to immediate follow-up, 12/30 to 6 months, and 7/30 to 12 months; only three received longer intervals, one each at 24, 36 and 48 months. Overall, 90.0% of patients were recommended for follow-up within 12 months. Clinical adjustment substantially changed image-only recommendations (Figure 1a), which were initially concentrated at 12 months, with no immediate follow-up assignments. After adjustment, most changes shifted patients toward shorter intervals: among those initially assigned to 12 months, 6 moved to immediate follow-up and 9 to 6 months, while 3 remained stable and 1 moved to 24 months. Patients initially assigned to 6 or 24 months showed more heterogeneous transitions, including both shorter and longer final intervals. Clinical contributions were patient-specific (Figure 1b). Diabetes duration showed the clearest positive contribution to risk, particularly in patients assigned to shorter follow-up, while HbA1c, systolic blood pressure and sex contributed in both directions. Fenofibrate had no relevant effect, and anaemia contributed only in few patients. Overall, final recommendations reflected the combination of image-derived risk and selected clinical modifiers.

**Figure 1:**
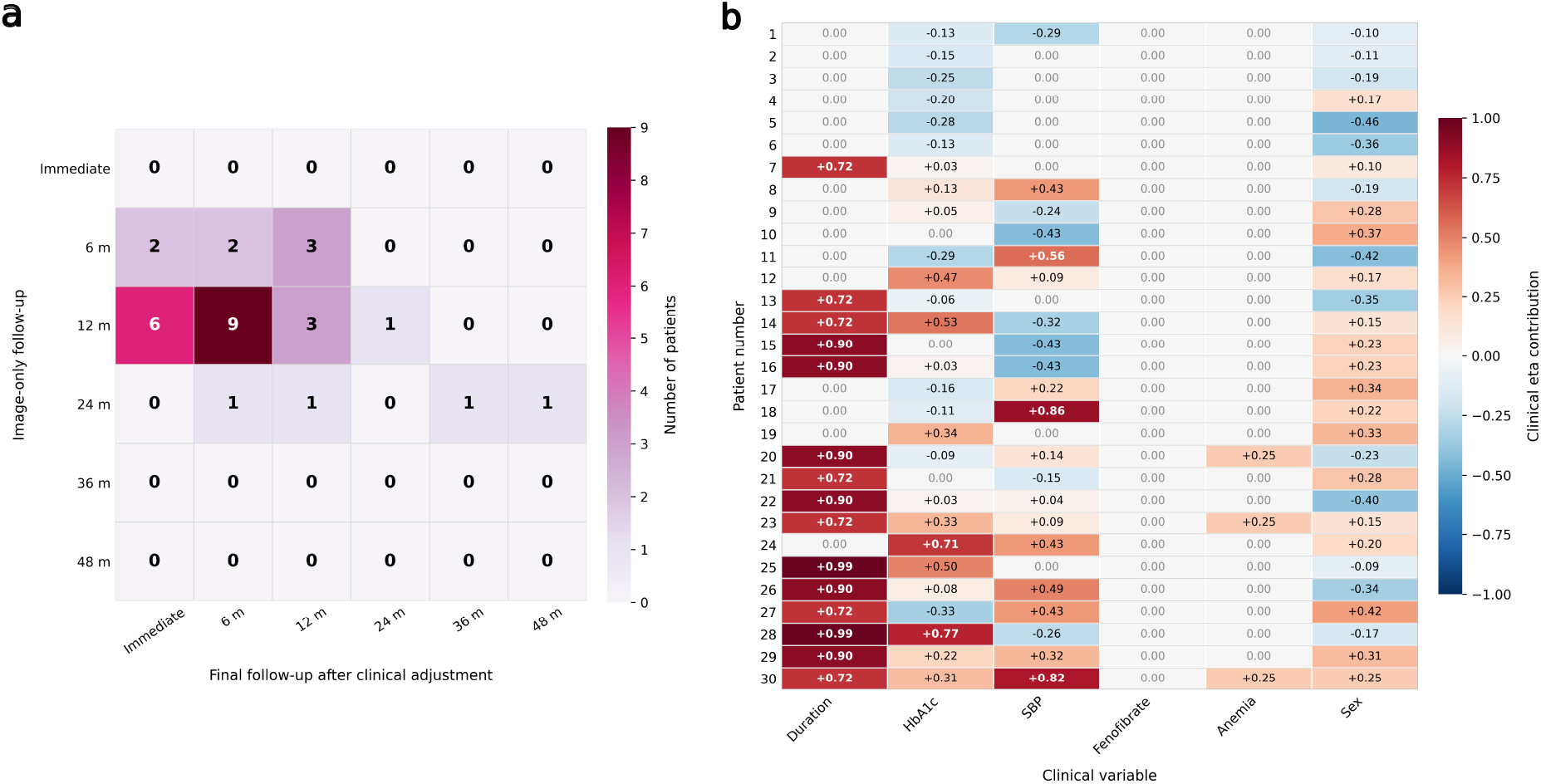
Application of the integrated image–clinical model for personalized diabetic retinopathy follow-up recommendations. Panel a, transition matrix comparing image-only follow-up recommendations with final clinically adjusted recommendations. Panel b, patient-specific contributions (*η* values) of individual clinical variables to the personalized risk adjustment model.

## 4 Conclusion

This study presents an integrated image–clinical framework for personalized diabetic retinopathy follow-up. By combining fundus image classification with literature-derived clinical risk modifiers, the model provides patient-specific progression risk estimates and shows that clinical adjustment can substantially modify image-only recommendations. Although preliminary and limited by the size of the target cohort, these findings support the feasibility of using an image–clinical risk model to inform personalized screening intervals in diabetic retinopathy. Further validation on larger, longitudinal, and clinically representative cohorts will be required to assess the safety of the proposed recommendations before real-world implementation.

## Data Availability

Datasets and code supporting this study are available from the authors upon reasonable request.

## Conflict of interests

The authors declare no conflict of interests.

## Funding

This research was funded by the “Digital Health and Artificial Intelligence” project, supported by the Autonomous Province of Trento (Provincial Government Resolution No. 2475, December 22, 2022).

## Availability of data and software code

Datasets and code supporting this study are available from the authors upon reasonable request.

## Notes

### Competing Interest Statement

The authors have declared no competing interest.

### Author Declarations

The study protocol was approved by the Territorial Ethics Committee of the Autonomous Province of Trento on 20 March 2024 (Ethics Committee register no. A958; protocol code INSPIRED-2023), and written informed consent was obtained from all participants prior to enrollment.

